# Development of a data-driven COVID-19 prognostication tool to inform triage and step-down care for hospitalised patients in Hong Kong: A population-based cohort study

**DOI:** 10.1101/2020.07.13.20152348

**Authors:** Eva L.H. Tsui, Carrie S.M. Lui, Pauline P.S. Woo, Alan T.L. Cheung, Peggo K.W. Lam, Van T.W. Tang, C.F. Yiu, C.H. Wan, Libby H.Y. Lee

**Affiliations:** Statistics and Data Science, Department, Hospital Authority, Hong Kong, Address: Hospital Authority Building, 147B Argyle Street, Kowloon, Hong Kong; Strategy and Planning Division, Hospital Authority, Hong Kong, Address: Hospital Authority Building, 147B Argyle Street, Kowloon, Hong Kong

**Keywords:** COVID-19, prognostic, prediction, clinical outcome, disease severity, triage, step-down care

## Abstract

**Background:** This is the first study on prognostication in an entire cohort of laboratory-confirmed COVID-19 patients in the city of Hong Kong. Prognostic tool is essential in the contingency response for the next wave of outbreak. This study aims to develop prognostic models to predict COVID-19 patients’ clinical outcome on day 1 and day 5 of hospital admission.

**Methods:** We did a retrospective analysis of a complete cohort of 1,037 COVID-19 laboratory-confirmed patients in Hong Kong as of 30 April 2020, who were admitted to 16 public hospitals with their data sourced from an integrated electronic health records system. It covered demographic information, chronic disease(s) history, presenting symptoms as well as the worst clinical condition status, biomarkers’ readings and Ct value of PCR tests on Day-1 and Day-5 of admission. The study subjects were randomly split into training and testing datasets in a 8:2 ratio. Extreme Gradient Boosting (XGBoost) model was used to classify the training data into three disease severity groups on Day-1 and Day-5.

**Results:** The 1,037 patients had a mean age of 37.8 (SD±17.8), 53.8% of them were male. They were grouped under three disease outcome: 4.8% critical/serious, 46.8% stable and 48.4% satisfactory. Under the full models, 30 indicators on Day-1 and Day-5 were used to predict the patients’ disease outcome and achieved an accuracy rate of 92.3% and 99.5%. With a trade-off between practical application and predictive accuracy, the full models were reduced into simpler models with seven common specific predictors, including the worst clinical condition status (4-level), age group, and five biomarkers, namely, CRP, LDH, platelet, neutrophil/lymphocyte ratio and albumin/globulin ratio. Day-1 model’s accuracy rate, macro- and micro-averaged sensitivity and specificity were 91.3%, 84.9%-91.3% and 96.0%-95.7% respectively, as compared to 94.2%, 95.9%-94.2% and 97.8%-97.1% under Day-5 model.

**Conclusions:** Both Day-1 and Day-5 models can accurately predict the disease severity. Relevant clinical management could be planned according to the predicted patients’ outcome. The model is transformed into a simple online calculator to provide convenient clinical reference tools at the point of care, with an aim to inform clinical decision on triage and step-down care.

## Introduction

The World Health Organization (WHO) had declared a pandemic outbreak of a new coronavirus, named COVID-19, on 12 March 2020. As of 30 April 2020, Hong Kong (HK) had a total of 1,037 confirmed cases as compared to a global caseload of over three million and mortality of over 210,000 at the same snapshot [1]. Four deaths [2] were reported locally. All patients were admitted to the Hospital Authority (HA) hospitals for management. HK has adopted the “early identification, early isolation and early treatment” infection control approach. She is ranked top to effectively curb COVID-19 transmission in the study comparing containment measures among different nations [3], given that HK has a population of 7.5 million residing in one of the most densely populated cities in the world [4].

HA is statutory, publicly funded organization to provide public medical care to all the citizens in HK. The services are organized via seven hospital clusters comprising of 43 hospitals, 49 specialist outpatient and 73 primary care clinics. It provides over 90% of hospital bed days in the territory through a total of 29,435 beds[5]. HA provides a single electronic health record system via the integrated Clinical Management System (CMS) [6], so that data of patients utilizing the service are automatically captured. Since the SARS epidemic in 2003, HA has made available over 1,200 airborne infection isolation (AII) beds across 16 public hospitals to support outbreak of infectious diseases. In view of the unprecedented rapidly growing number of infections in the neighbouring Mainland China in mid-February 2020, the HA management implemented a proactive response plan to convert some acute general wards into “retrofit wards” [7] of negative pressures and directed airflow.

In pursuit of the early identification strategy, Hong Kong has been incrementally widening the surveillance coverage to identify and contact trace suspected cases through various channels. All suspected cases are referred to the HA for management. Samples will be sent to the public laboratory in the Department of Health for confirmation: two positive consecutive reverse-transcription polymerase chain reaction (RT-PCR) tests [8, 9] are required, as suggested by the WHO [10]. The patients can only be discharged from the hospital care when the isolation order is lift, given that their clinical conditions improve and they are afebrile with two negative RT-PCR results taken at least 24 hours apart [8, 9].

Starting from mid-March, there was second wave with a surge in imported cases from overseas via airport arrivals. In early April, the occupancy rate of our AII rooms and beds peaked at 78% and 70% respectively, which was approaching saturation. The demand pressure on first-tier isolation beds could be partly relieved by the timely conversion and operation of some 400 second-tier isolation beds in 10 hospitals. Patients with stable clinical features but yet to fulfill the discharge criteria are transferred to the second-tier ward for further management. A prognostic model was developed with an aim to early identify those cases with satisfactory or stable clinical outcome for step-down care.

The outbreak in HK slows down since the end of April, with 1,047 confirmed cases as of 10 May, which is the cut-off date of data analytics for this study. However, learning the experience from nearby countries like Singapore, HK remains vigilant and is preparing for the possible third wave of infection once the international travels resume normal. The research team repeated the prognostic model based on the entire COVID-19 cohort, aiming to predict their clinical outcome as early as day 1 of admission.

## Methods

### Study population, data collection and definition

All 1,037 confirmed cases as of 30 April 2020 were included in this study, and their observational data were traced up to 10 May. Data sources come from internal systems, CMS and eNID of NDORS. “NDORS” is an electronic platform for both HA and the Department of Health, to digitally report all suspected and confirmed statutory notifiable diseases and other infectious diseases of public health concern. A designated “eNID” module for reporting COVID-19 cases and clinical management was specifically built and interfaced to NDORS [8, 9]. This study was a retrospective data analysis on de-identified patient-based electronic medical records from CMS and eNID. Person-based information on history of chronic diseases was retrieved from an established HA’s chronic disease virtual registry that contains 25 pre-defined chronic diseases (Table 1). The registry was electronically built and based on all past CMS’ medical records using some operational counting and classification rules specific to each non-cancer disease, together with cancer cases sourced from Hong Kong’s Cancer Registry [11].

**Table 1:**
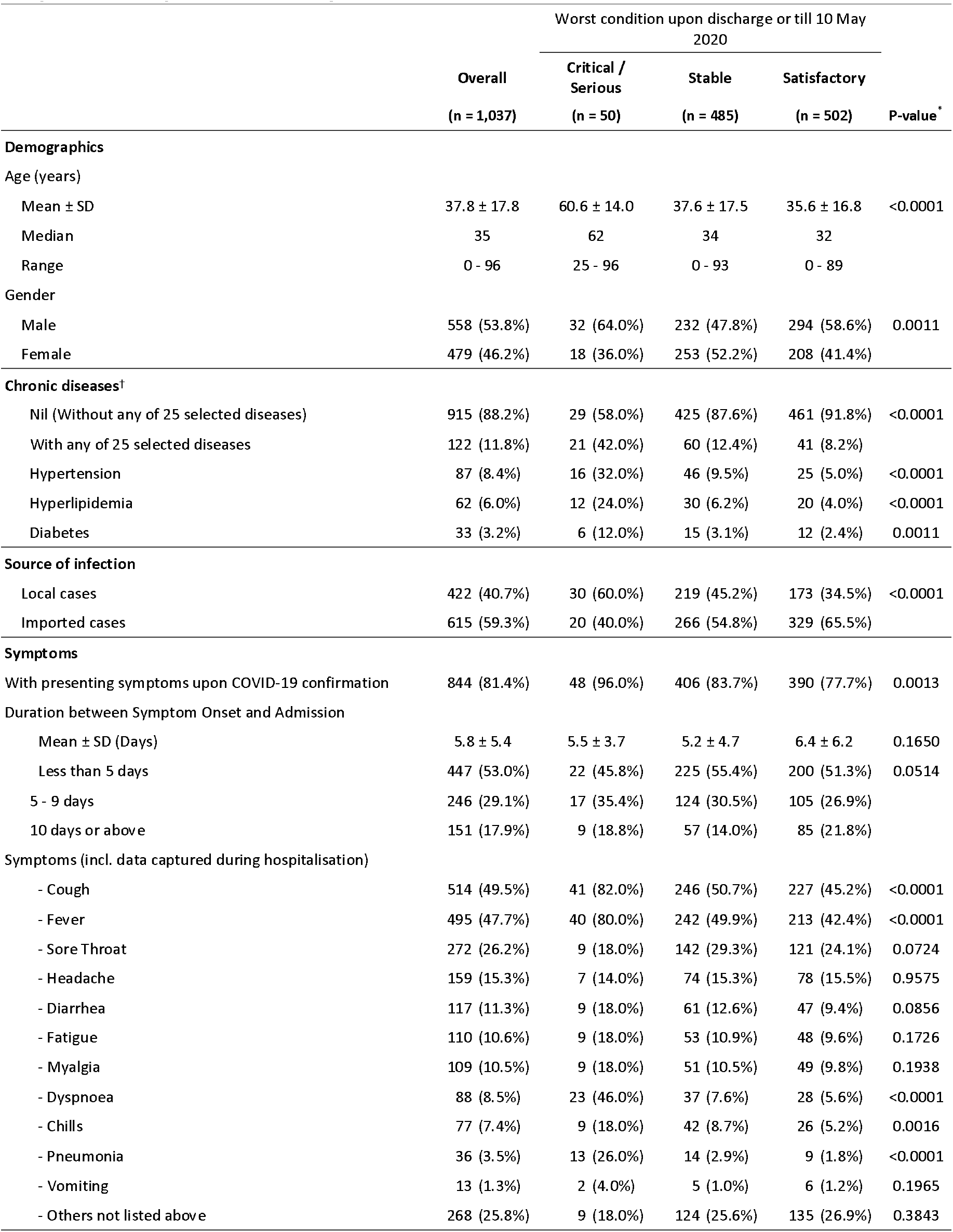
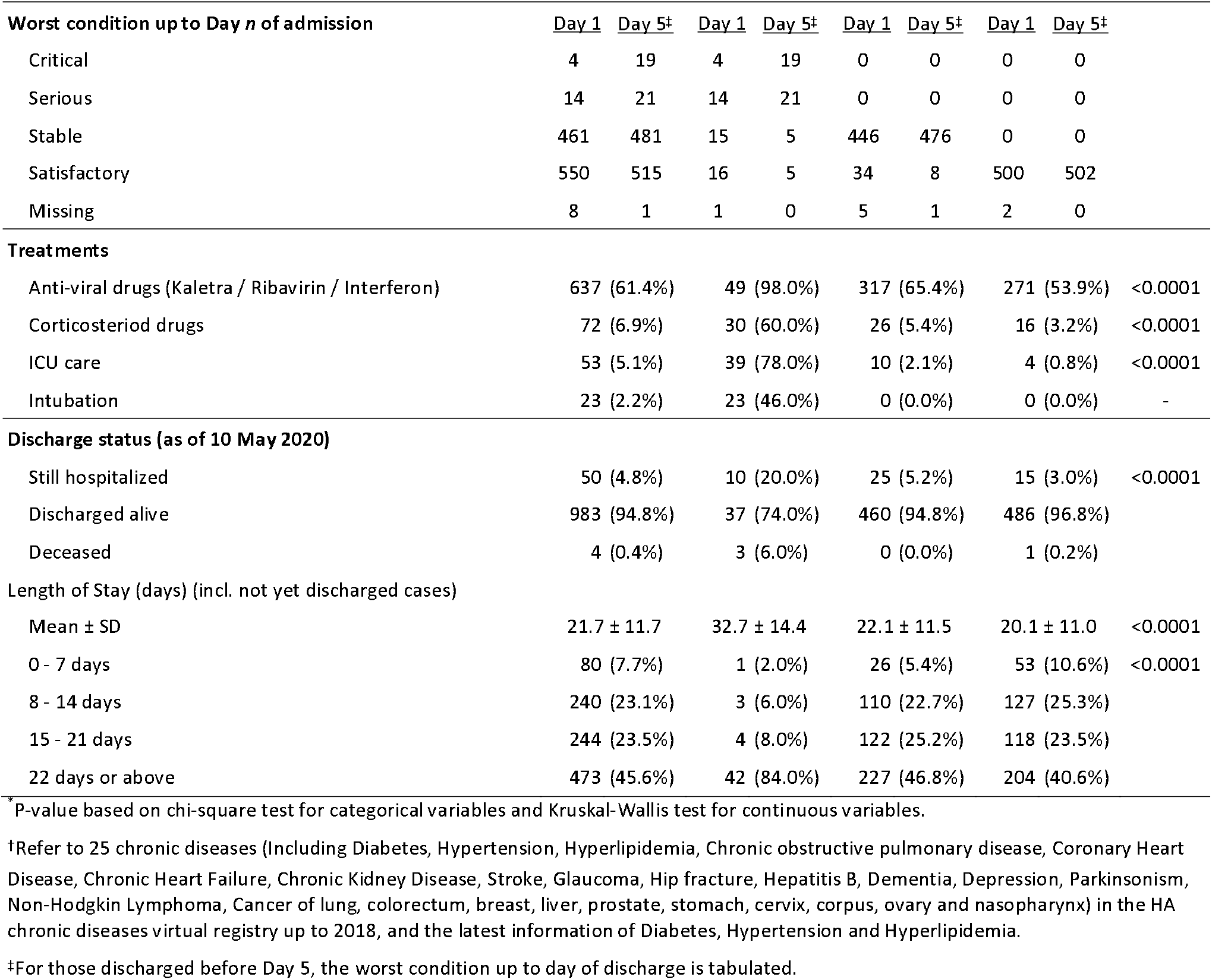
Patient profile of 1,037 COVID-19 confirmed cases as of 30 April 2020 (with data during hospitalisation updated till 10 May 2020)

|  | Worst condition upon discharge or till 10 May 2020 |  |  |  | P-value<sup>*</sup> |
| --- | --- | --- | --- | --- | --- |
|  | Overall | Critical / Serious | Stable | Satisfactory |  |
|  | (n = 1,037) | (n = 50) | (n = 485) | (n = 502) |  |
| Demographics |  |  |  |  |  |
| Age (years) |  |  |  |  |  |
| Mean ± SD | 37.8 ± 17.8 | 60.6 ± 14.0 | 37.6 ± 17.5 | 35.6 ± 16.8 | <0.0001 |
| Median | 35 | 62 | 34 | 32 |  |
| Range | 0 - 96 | 25 - 96 | 0 - 93 | 0 - 89 |  |
| Gender |  |  |  |  |  |
| Male | 558 (53.8%) | 32 (64.0%) | 232 (47.8%) | 294 (58.6%) | 0.0011 |
| Female | 479 (46.2%) | 18 (36.0%) | 253 (52.2%) | 208 (41.4%) |  |
| Chronic diseases<sup>†</sup> |  |  |  |  |  |
| Nil (Without any of 25 selected diseases) | 915 (88.2%) | 29 (58.0%) | 425 (87.6%) | 461 (91.8%) | <0.0001 |
| With any of 25 selected diseases | 122 (11.8%) | 21 (42.0%) | 60 (12.4%) | 41 (8.2%) |  |
| Hypertension | 87 (8.4%) | 16 (32.0%) | 46 (9.5%) | 25 (5.0%) | <0.0001 |
| Hyperlipidemia | 62 (6.0%) | 12 (24.0%) | 30 (6.2%) | 20 (4.0%) | <0.0001 |
| Diabetes | 33 (3.2%) | 6 (12.0%) | 15 (3.1%) | 12 (2.4%) | 0.0011 |
| Source of infection |  |  |  |  |  |
| Local cases | 422 (40.7%) | 30 (60.0%) | 219 (45.2%) | 173 (34.5%) | <0.0001 |
| Imported cases | 615 (59.3%) | 20 (40.0%) | 266 (54.8%) | 329 (65.5%) |  |
| Symptoms |  |  |  |  |  |
| With presenting symptoms upon COVID-19 confirmation | 844 (81.4%) | 48 (96.0%) | 406 (83.7%) | 390 (77.7%) | 0.0013 |
| Duration between Symptom Onset and Admission |  |  |  |  |  |
| Mean ± SD (Days) | 5.8 ± 5.4 | 5.5 ± 3.7 | 5.2 ± 4.7 | 6.4 ± 6.2 | 0.1650 |
| Less than 5 days | 447 (53.0%) | 22 (45.8%) | 225 (55.4%) | 200 (51.3%) | 0.0514 |
| 5 - 9 days | 246 (29.1%) | 17 (35.4%) | 124 (30.5%) | 105 (26.9%) |  |
| 10 days or above | 151 (17.9%) | 9 (18.8%) | 57 (14.0%) | 85 (21.8%) |  |
| Symptoms (incl. data captured during hospitalisation) |  |  |  |  |  |
| - Cough | 514 (49.5%) | 41 (82.0%) | 246 (50.7%) | 227 (45.2%) | <0.0001 |
| - Fever | 495 (47.7%) | 40 (80.0%) | 242 (49.9%) | 213 (42.4%) | <0.0001 |
| - Sore Throat | 272 (26.2%) | 9 (18.0%) | 142 (29.3%) | 121 (24.1%) | 0.0724 |
| - Headache | 159 (15.3%) | 7 (14.0%) | 74 (15.3%) | 78 (15.5%) | 0.9575 |
| - Diarrhea | 117 (11.3%) | 9 (18.0%) | 61 (12.6%) | 47 (9.4%) | 0.0856 |
| - Fatigue | 110 (10.6%) | 9 (18.0%) | 53 (10.9%) | 48 (9.6%) | 0.1726 |
| - Myalgia | 109 (10.5%) | 9 (18.0%) | 51 (10.5%) | 49 (9.8%) | 0.1938 |
| - Dyspnoea | 88 (8.5%) | 23 (46.0%) | 37 (7.6%) | 28 (5.6%) | <0.0001 |
| - Chills | 77 (7.4%) | 9 (18.0%) | 42 (8.7%) | 26 (5.2%) | 0.0016 |
| - Pneumonia | 36 (3.5%) | 13 (26.0%) | 14 (2.9%) | 9 (1.8%) | <0.0001 |
| - Vomiting | 13 (1.3%) | 2 (4.0%) | 5 (1.0%) | 6 (1.2%) | 0.1965 |
| - Others not listed above | 268 (25.8%) | 9 (18.0%) | 124 (25.6%) | 135 (26.9%) | 0.3843 |

**Table 1: Patient profile of 1,037 COVID-19 confirmed cases as of 30 April 2020 (with data during hospitalisation updated till 10 May 2020) (Cont'd)**
|  | Worst condition upon discharge or till 10 May 2020 |  |  |  |  |  |  |  | P-value<sup>*</sup> |
| --- | --- | --- | --- | --- | --- | --- | --- | --- | --- |
|  | Overall |  | Critical / Serious |  | Stable |  | Satisfactory |  |  |
|  | (n = 1,037) |  | (n = 50) |  | (n = 485) |  | (n = 502) |  |  |
| Worst condition up to Day *n* of admission | Day 1 | Day 5<sup>‡</sup> | Day 1 | Day 5<sup>‡</sup> | Day 1 | Day 5<sup>‡</sup> | Day 1 | Day 5<sup>‡</sup> |  |
| Critical | 4 | 19 | 4 | 19 | 0 | 0 | 0 | 0 |  |
| Serious | 14 | 21 | 14 | 21 | 0 | 0 | 0 | 0 |  |
| Stable | 461 | 481 | 15 | 5 | 446 | 476 | 0 | 0 |  |
| Satisfactory | 550 | 515 | 16 | 5 | 34 | 8 | 500 | 502 |  |
| Missing | 8 | 1 | 1 | 0 | 5 | 1 | 2 | 0 |  |
| Treatments |  |  |  |  |  |  |  |  |  |
| Anti-viral drugs (Kaletra / Ribavirin / Interferon) | 637 (61.4%) |  | 49 (98.0%) |  | 317 (65.4%) |  | 271 (53.9%) |  | <0.0001 |
| Corticosteriod drugs | 72 (6.9%) |  | 30 (60.0%) |  | 26 (5.4%) |  | 16 (3.2%) |  | <0.0001 |
| ICU care | 53 (5.1%) |  | 39 (78.0%) |  | 10 (2.1%) |  | 4 (0.8%) |  | <0.0001 |
| Intubation | 23 (2.2%) |  | 23 (46.0%) |  | 0 (0.0%) |  | 0 (0.0%) |  | - |
| Discharge status (as of 10 May 2020) |  |  |  |  |  |  |  |  |  |
| Still hospitalized | 50 (4.8%) |  | 10 (20.0%) |  | 25 (5.2%) |  | 15 (3.0%) |  | <0.0001 |
| Discharged alive | 983 (94.8%) |  | 37 (74.0%) |  | 460 (94.8%) |  | 486 (96.8%) |  |  |
| Deceased | 4 (0.4%) |  | 3 (6.0%) |  | 0 (0.0%) |  | 1 (0.2%) |  |  |
| Length of Stay (days) (incl. not yet discharged cases) |  |  |  |  |  |  |  |  |  |
| Mean ± SD | 21.7 ± 11.7 |  | 32.7 ± 14.4 |  | 22.1 ± 11.5 |  | 20.1 ± 11.0 |  | <0.0001 |
| 0 - 7 days | 80 (7.7%) |  | 1 (2.0%) |  | 26 (5.4%) |  | 53 (10.6%) |  | <0.0001 |
| 8 - 14 days | 240 (23.1%) |  | 3 (6.0%) |  | 110 (22.7%) |  | 127 (25.3%) |  |  |
| 15 - 21 days | 244 (23.5%) |  | 4 (8.0%) |  | 122 (25.2%) |  | 118 (23.5%) |  |  |
| 22 days or above | 473 (45.6%) |  | 42 (84.0%) |  | 227 (46.8%) |  | 204 (40.6%) |  |  |

<sup>\*</sup>P-value based on chi-square test for categorical variables and Kruskal-Wallis test for continuous variables.
<sup>†</sup>Refer to 25 chronic diseases (Including Diabetes, Hypertension, Hyperlipidemia, Chronic obstructive pulmonary disease, Coronary Heart Disease, Chronic Heart Failure, Chronic Kidney Disease, Stroke, Glaucoma, Hip fracture, Hepatitis B, Dementia, Depression, Parkinsonism, Non-Hodgkin Lymphoma, Cancer of lung, colorectum, breast, liver, prostate, stomach, cervix, corpus, ovary and nasopharynx) in the HA chronic diseases virtual registry up to 2018, and the latest information of Diabetes, Hypertension and Hyperlipidemia.
<sup>‡</sup>For those discharged before Day 5, the worst condition up to day of discharge is tabulated.

All 16 HA hospitals that treated COIVD-19 cases adopted a unified classification scheme on clinical conditions. The in-charge physicians would continuously update the condition status whenever the patient deteriorated or improved. The four clinical conditions are (1) *critical*: require intubation, or extracorporeal membrane oxygenation (ECMO) or in shock; (2) *serious*: require oxygen supplement of 3 litres or more per minute; (3) *stable*: with mild influenza-like illness (ILI) symptoms; (4) *satisfactory*: progressing well and likely to be discharged soon. Based on their clinical condition(s) along the entire clinical course, all cases were further amalgamated into three distinct outcome groups to delineate a grading for disease severity. The groups are “critical/serious”, “stable” and “satisfactory”. The patients must have ever been assessed as either “critical” or “serious” clinical condition for one or more days in the “critical/serious” group; otherwise being classified into the second group if ever assessed as “stable”. The remaining third group must be entirely assessed as “satisfactory” along the clinical course.

### Statistical analysis

Descriptive statistical analyses were performed for the entire cohort with respect to epidemiological, clinical and laboratory data. In addition, chi-square test for categorical variables and Kruskal-Wallis test for continuous variables were performed to evaluate if there were any differences in a host of prognostic factors on day 1 and day 5 of hospital admission among the three outcome groups. For development as well as evaluation of the model, the entire 1,037 study subjects were, proportional to outcome distribution, randomly split into a training dataset comprising of 829 subjects and a testing dataset of the remaining 208 subjects. The Extreme Gradient Boosting (XGBoost) model, which is a boosting decision tree machine learning framework allowing missing values for individual predictor variables, was developed to classify the training data into one of the three outcome groups, after taking into account a host of 30 predictors which included age, gender, chronic disease(s) history, 11 presenting symptoms as well as the worst clinical condition status, 15 biomarkers’ readings and Ct value of RT-PCR tests (based on E-Gene of the TIB MIBIOL kit) on day 1 and day 5 of admission. These predictors were chosen with reference to studies on COVID-19 [12–23] and SARS [24–26].

The XGBoost classifiers were trained and tuned using a 5-fold cross-validation approach with the training data to obtain the optimal hyperparameters [27]. All 30 features were ranked according to their relative importance using F-score, which guided a variable selection process to reduce the full model into a simpler one for practical application. The model output for each subject a probability across each of the three outcome groups, summing up to one, and with the highest probability group as the predicted outcome class. After applying the trained model to the testing dataset, it was then analysed in a 3×3 confusion matrix, based on which the model’s overall accuracy rate was computed. In view of the imbalanced outcome group distribution, macro-averaged and micro-averaged sensitivity and specificity of the three outcome groups were derived to evaluate the model performance. Decision tree of the simplified classifiers was also output for each outcome group. Partial dependency plots were output to depict the marginal effect of each model feature on the predicted outcome. (Appendix Figure 1) The XGBoost models were carried out by using Python’s XGboost version 1.10 whereas other statistical analyses by SAS version 9.4 software.

## Results

### Epidemiological and clinical profile

Table 1 provides a complete epidemiological and clinical profile of all 1,037 laboratory confirmed COVID-19 cases, 95.2% had been discharged (including 4 deceased cases) and the remaining 50 (4.8%) cases have stayed for 19-70 days as at data analytics cut-off date. All the subjects were post-stratified into three pre-defined outcome groups: 50 (4.8%) in the critical/serious group, 485 (46.8%) stable group and 502 (48.4%) satisfactory group, which can then be compared against their respective clinical condition (4-level) on day 1 and day 5 of admission.

The study subjects were aged from 1 month to 96 years. Critical/serious group was significantly older (mean 60.6 years, SD±14.0) than stable (mean 37.6 years, SD±17.5) and satisfactory (mean 35.6 years, SD±16.8) group (p<0.0001). Over half (53.8%) of total caseload was male, comparatively higher in critical/serious group (64.0%) than stable (47.8%) and satisfactory (58.6%) groups (p=0.001).

Chronic disease(s) were present in 11.8% of this study cohort, with most common ones being hypertension, hyperlipidemia and diabetes. A significant difference (p<0.0001) in chronic disease(s) prevalence was found: 42.0%, 12.4% and 8.2% in critical / serious, stable and satisfactory group respectively.

81.4% of all the cases were symptomatic upon COVID-19 confirmation. Cough and fever were the top two symptoms prevalent in almost half of total confirmed cases, whereas sore throat in around one-quarter. Among the symptomatic cases, the mean duration between symptom onset and admission was 5.8 days (SD±5.4). Comparatively, the satisfactory outcome group had a significantly (p<0.0001) longer duration (6.4 days, SD±6.2) than the stable and critical/serious groups’ (5.2 days, SD±4.7; 5.5 days, SD±3.7).

Table 1 shows the proportion of patients on anti-viral and corticosteroid drugs. ICU care and intubation treatments were respectively provided to 78.0% and 46.0% in critical/serious outcome groups.

### Laboratory readings on Day 1 and Day 5 of admission

This study defines day 0 according to the time of admission recorded at admission offices from 00:00 to 23:59. A higher proportion of missing values on day 0 is expected because three-quarters of the study subjects had admitted for less than 12 hours. Therefore, Table 2 shows the latest laboratory readings as of day 1 and day 5. For those tests whose normal reference ranges vary across HA laboratories, they are expressed as multiples of the upper normal reference.

**Table 2:**
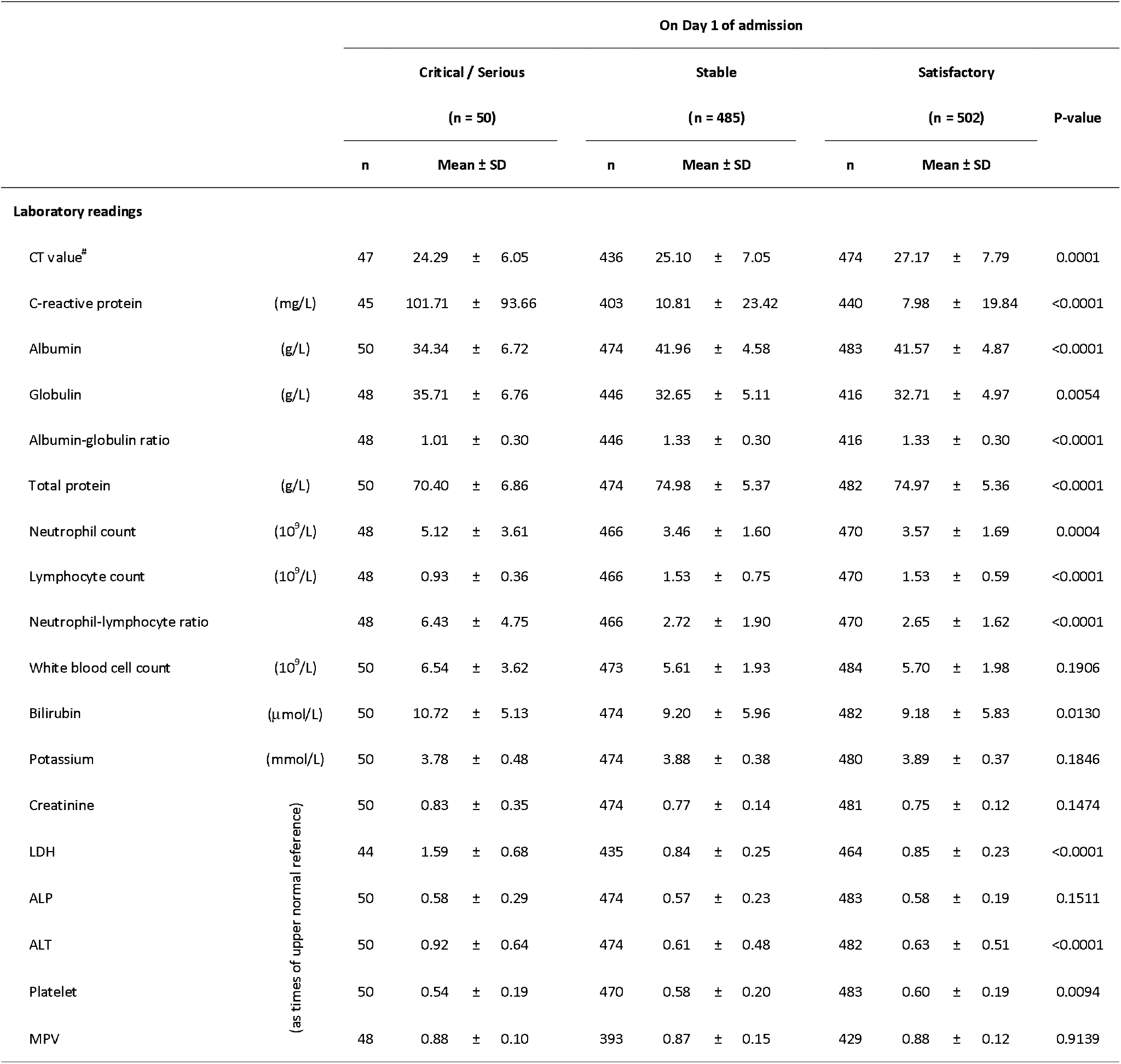
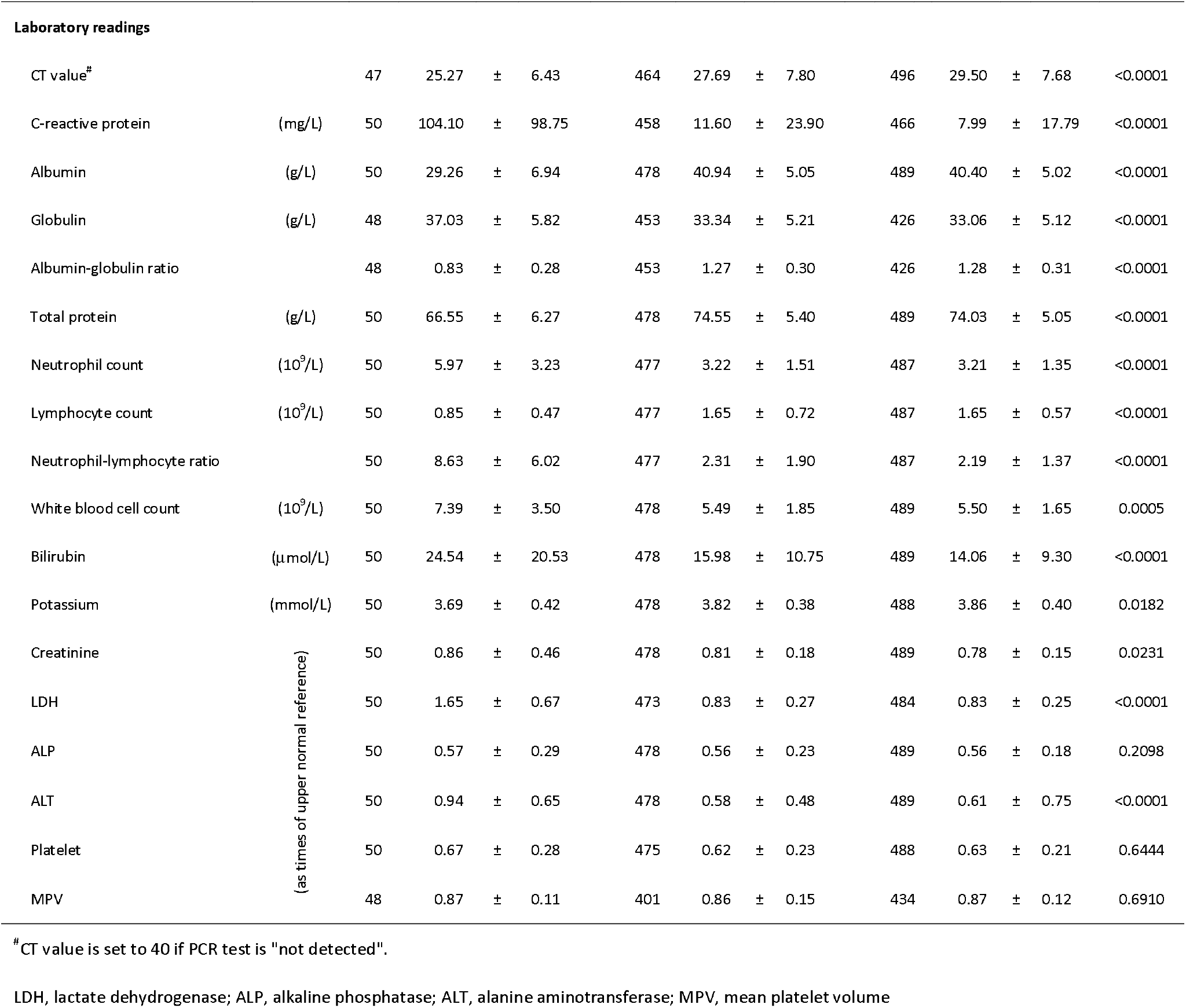
Laboratory readings on Day 1 and Day 5 of admission among 1,037 COVID-19 confirmed cases as of 30 April 2020.

Across the 15 biomarkers and two derived ratios (albumin-globulin ratio, neutrophil-lymphocyte ratio), no statistically significant between-group differences were found in mean platelet volume (MPV) and alkaline phosphatase (ALP) on both days 1 and 5, white blood count cell (WBC) on day 1 whereas platelet on day 5. Otherwise, statistical significance was stronger on day 5 than day 1 for all other tests except lactate dehydrogenase (LDH) which had a reverse pattern. Five biomarkers had strongest statistical significance (p<0.0001) in terms of between-group differences on both days, namely C-reactive protein (CRP), lymphocyte counts, albumin (A), globulin (G) and total protein tests. Their respective readings in terms of mean and standard deviation across the three outcome groups are shown in Table 2. Based on the same blood specimen, N/L and A/G ratios can amalgamate and amplify the effects of each pair of biomarkers whose scale in opposite direction, contributed to a much stronger discriminatory effect on both days (p<0.0001).

Ct value of RT-PCR tests for E-Gene of the TIB MIBIOL kit indicates the viral load, with low value representing high viral load and a theoretical maximum of 40. Significant between-group differences in Ct value were found, on day 1 with lowest mean reading in critical/serious group (24.3±6.1), followed by stable group (25.1±7.1) and then satisfactory group (27.2±7.8) (p=0.0001); and their corresponding values on day 5 were 25.3±6.4, 27.7±7.8 and 29.5±7.7 (p<0.0001).

### Prediction model performance and application

#### The full model

For the XGBoost decision tree model on day 1 and day 5, all 30 predictors were ranked according to their relative importance based on F-score in Figure 1. Based on the testing dataset (n=208), an extremely strong concordance between the predicted and actual outcome classification in the confusion matrix was found for both models. The overall accuracy rate of the Day-1 model was 92.3%, with 82.6%-92.3% macro- and micro-averaged sensitivity and 96.0%-96.1% macro- and micro-averaged specificity. As for the Day-5 model, the overall accuracy rate was 99.5%, with the corresponding sensitivity at 99.5%-99.7% and specificity at 99.5%. (Table 3)

**Figure 1:**
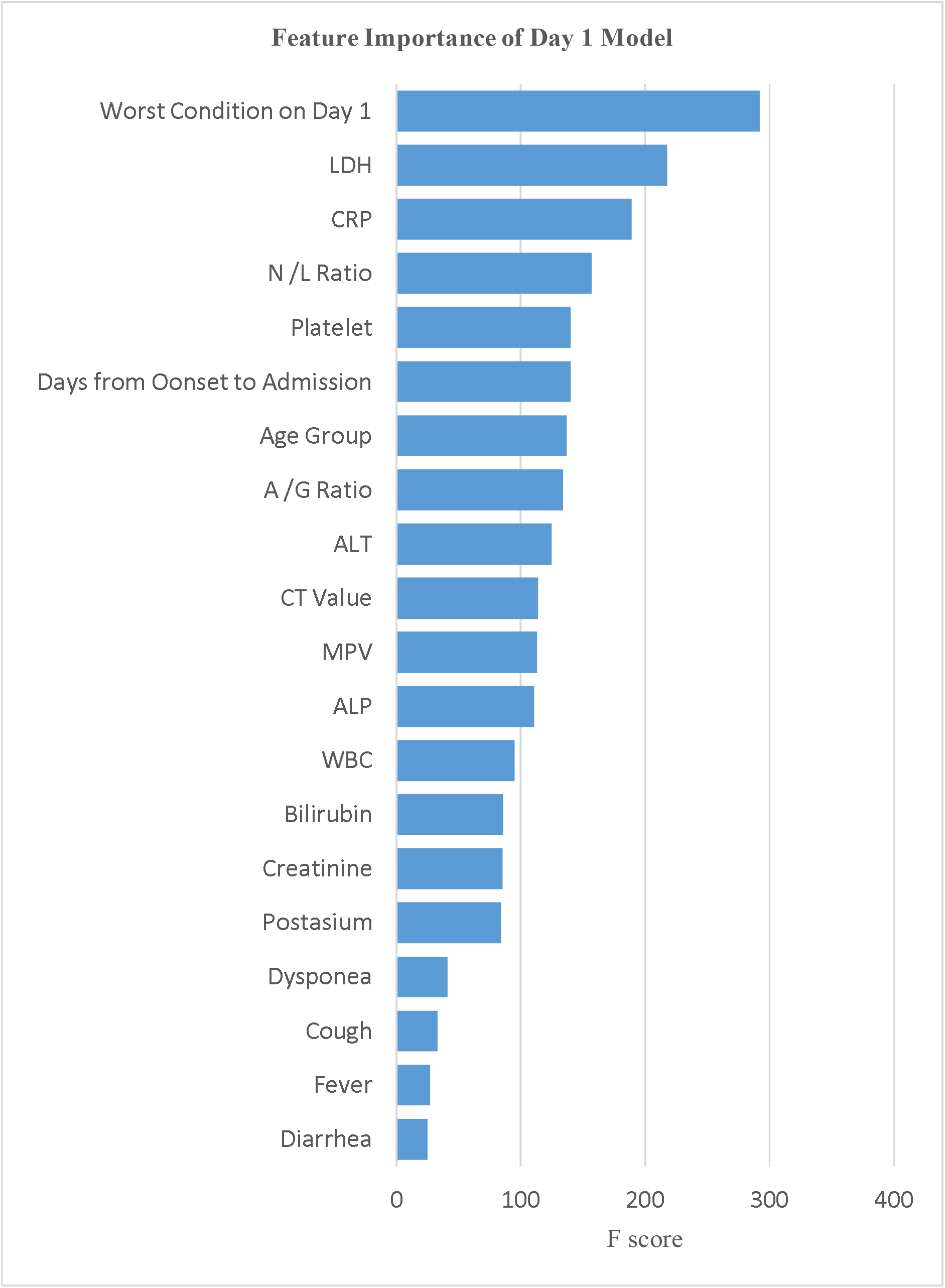
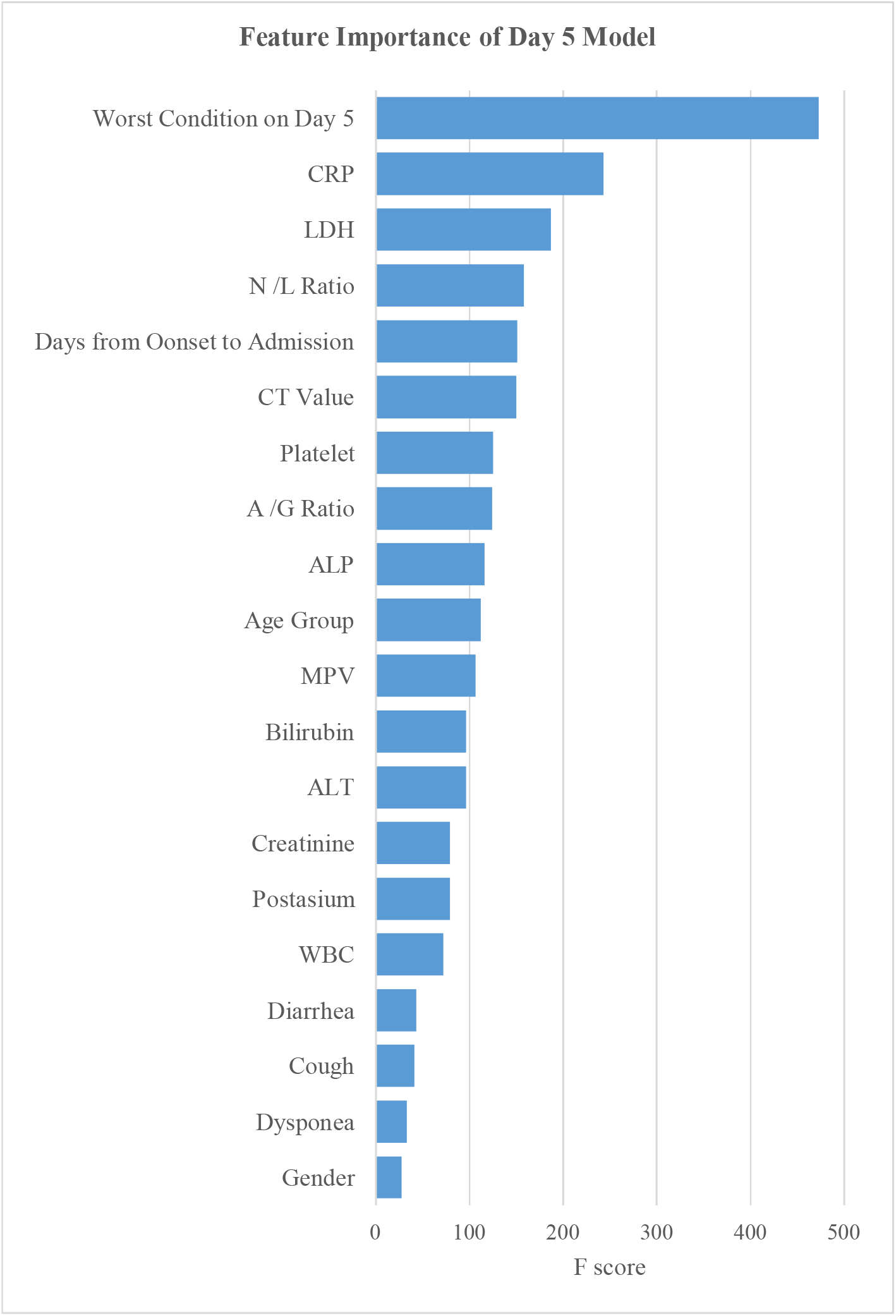
Top 20 Features* ranked according to importance in the XGBoost model. *Feature importance of total protein, gender, pneumonia, chronic disease, sore throat, fatigue, myalgia, chills, headache, and vomiting under Day-1 model and that of total protein, myalgia, pneumonia, fever, vomiting, chronic disease, headache, fatigue, sore throat, chills under Day-5 model were excluded from this figure

**Table 3:** Predictive performance of the full model and the simplified model.

|  |  | Full Model* (based on 30 features) |  |  |  |  |  |
| --- | --- | --- | --- | --- | --- | --- | --- |
|  |  | On Day 1 of admission |  |  | On Day 5 of admission<sup>#</sup> |  |  |
|  |  | Predicted Class |  |  | Predicted Class |  |  |
|  |  | Critical /<br>Serious | Stable | Satisfactory | Critical /<br>Serious | Stable | Satisfactory |
| Actual Class | Critical / Serious | 6 | 0 | 4 | 10 | 0 | 0 |
|  | Stable | 6 | 86 | 5 | 0 | 96 | 1 |
|  | Satisfactory | 1 | 0 | 100 | 0 | 0 | 101 |
| Sensitivity | Macro averaged | 82.6% |  |  | 99.7% |  |  |
|  | Micro averaged | 92.3% |  |  | 99.5% |  |  |
| Specificity | Macro averaged | 96.0% |  |  | 99.5% |  |  |
|  | Micro averaged | 96.1% |  |  | 99.5% |  |  |
| Accuracy |  | 92.3% |  |  | 99.5% |  |  |

|  |  | Simplified Model* (based on 7 features) |  |  |  |  |  |
| --- | --- | --- | --- | --- | --- | --- | --- |
|  |  | On Day 1 of admission |  |  | On Day 5 of admission<sup>#</sup> |  |  |
|  |  | Predicted Class |  |  | Predicted Class |  |  |
|  |  | Critical /<br>Serious | Stable | Satisfactory | Critical /<br>Serious | Stable | Satisfactory |
| Actual Class | Critical / Serious | 7 | 1 | 2 | 10 | 0 | 0 |
|  | Stable | 7 | 86 | 4 | 9 | 87 | 1 |
|  | Satisfactory | 4 | 0 | 97 | 2 | 0 | 99 |
| Sensitivity | Macro averaged | 84.9% |  |  | 95.9% |  |  |
|  | Micro averaged | 91.3% |  |  | 94.2% |  |  |
| Specificity | Macro averaged | 96.0% |  |  | 97.8% |  |  |
|  | Micro averaged | 95.7% |  |  | 97.1% |  |  |
| Accuracy |  | 91.3% |  |  | 94.2% |  |  |

\*Model performance based on testing dataset (n = 208)
<sup>#</sup>upon discharge if hospital discharged before Day 5

#### The simplified model associated with a calculator tool

With reference to their relative importance as indicated by F-scores, the next step was to optimize the selection of a smaller set of variables for training alternative simpler models at an opportunity cost of a reduction in accuracy rate. As a result, of the top 10 important features under Day-1 and Day-5 models (Figure 1), the same seven model features were selected and incorporated into two simplified models. They included the worst clinical condition status (4-level), age group, and five biomarkers, namely, CRP, LDH, platelet, N/L ratio and A/G ratio. Their discriminating effect across the three outcome groups is graphically shown in Appendix Figure 1. Onset-to-admission duration and Ct value of RT-PCR test were not selected due to concern of recall bias and limited applicability at other settings respectively. The alanine aminotransferase (ALT) and ALP biomarkers were also dropped as they were relatively less important and only appeared in either one model. The overall accuracy rate of this simplified Day-1 model was 91.3%, with macro- and micro-averaged sensitivity and specificity respectively at 84.9%-91.3% and 95.7%-96.0%. For the Day-5 model, the overall accuracy rate was 94.2%, with the corresponding sensitivity at 95.9%-94.2% and specificity at 97.1%-97.8%. (Table 3) The decision trees for each outcome group of the Day-1 simplified model are visualized in Figure 2.

**Figure 2:**
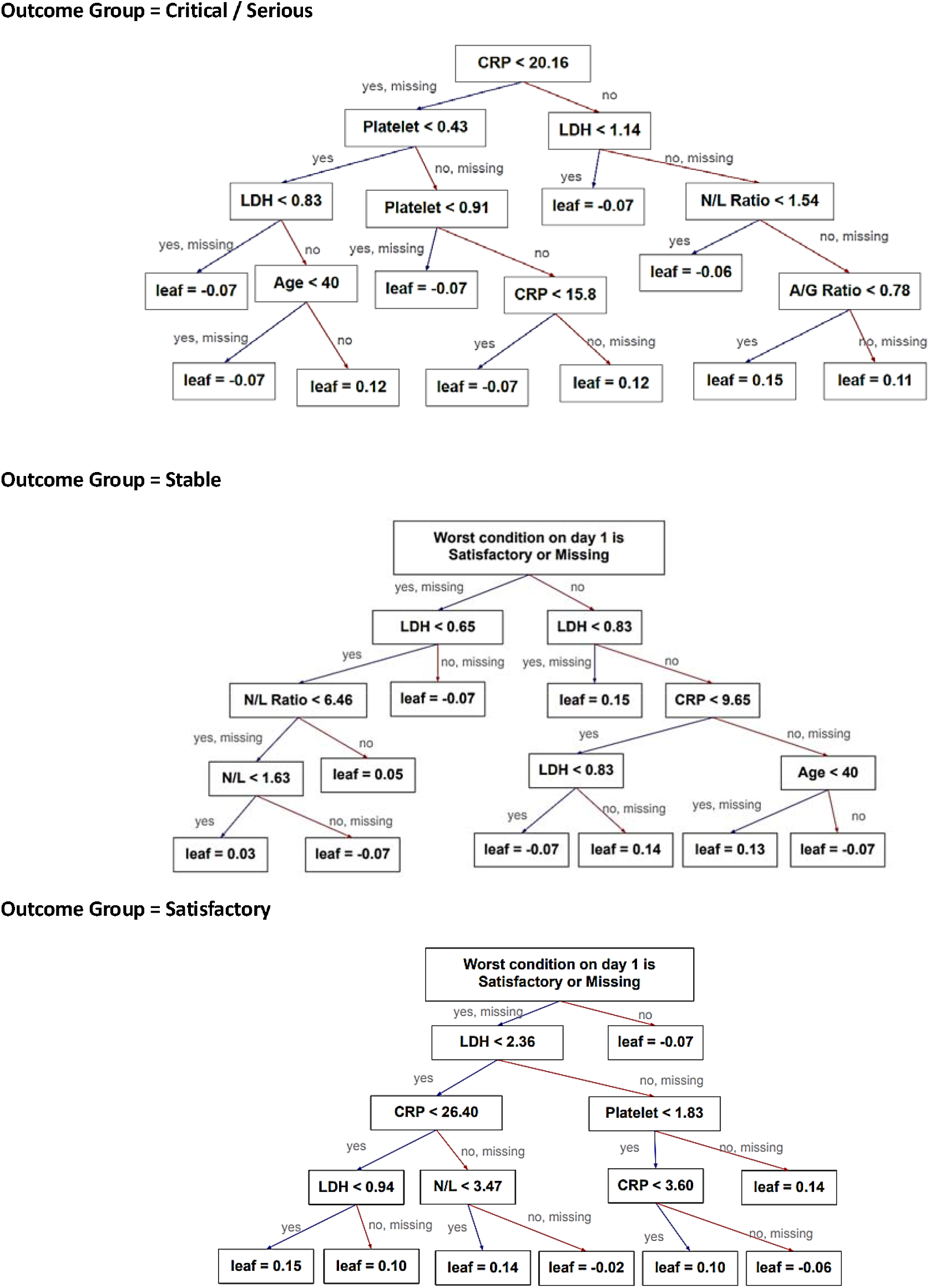
Decision rules using the key features under the Day-1 simplified model and their thresholds.

From Table 1, among the 32 patients who deteriorated from satisfactory or stable to critical/serious condition after day 1, the simplified Day-1 model has correctly predicted outcome group for 22 of them (4 out of 7 in the testing data and 18 out of 25 in the training data). For another 10 patients who deteriorated after day 5, all of them can be correctly classified by the Day-5 model.

## Discussion

The aim of the study is to provide additional information for early clinical decision-making in transferring COVID-19 patients to step-down care. This model utilizes the demographic, presenting symptoms, clinical and laboratory findings of the entire Hong Kong cohort to provide the best available analysis for developing the prognostic models.

It is suggested that HK was facing a different cohort of COVID-19 patients as compared to China. As compared to Zhang’s study [13] on disease severity of 663 patients in Wuhan (14% critical, 48% severe, 38% moderate, 0.5% mild), HK had a totally reverse pattern with only 5% critical/serious cases (Table 1) with regard to similar clinical criteria defining “critical” and “serious” between China [13, 28] and HK. This huge difference can be partly explained by a younger age profile in Hong Kong’s entire cohort relative to Zhang’s study cohort (mean age of 38 years versus 56 years).

Most studies predicted fatal outcome of mortality [12–14, 17, 19, 21] and critical illness [17, 19, 20] of COVID-19 cases, except two studies in China [13, 18] on risk factors for disease severity spectrum. Since the mortality of HK COVID-19 cases is low (four deaths among 1,037 patients), the above predicted fatal outcome is not applicable to stratify the needs of our patients nor to inform subsequent management decision for step-down care. Local study is necessary to provide additional insight to the disease management.

Our study finds significant correlation of patients’ predisposing risk factors like older age, male sex and presence of chronic diseases, in particular those related to cardiovascular diseases, with adverse outcome in univariate analysis. Similar findings were reported in other studies [12, 13, 16, 17, 20, 22]. The presenting symptoms of dyspnoea and fatigue were significant indicator for poorer outcome in other COVID-19 studies [12, 13, 17, 20]; whereas only dyspnoea was found significant in univariate analysis but relatively less important as compared to biomarkers in this study.

For the biomarkers (individually or in ratios) in the top 20 important model features list (Figure 1), they could predict disease severity; and most of them are also reported as independent prognostic factors of COVID-19 in other studies (13,17-22). We did not include aspartate aminotransferase (AST), procalicitonin (PCT) and D-dimer as in other COVID-19 studies [12–14, 17, 22] because only 16%-23% of our study subjects had such biomarkers tested. A few important prognosticators of this study are also in common with three previous local studies on prognostication in SARS patients, in which elevated CRP, LDH, neutrophil count, ALT, creatinine and platelet counts predictive of mortality, ICU care or oxygenation failure (24–26).

The study showed that clinical conditions on day 1 and day 5 could predict the subsequent clinical outcome, particularly in the stable and satisfactory patient groups in addition to their biomarkers’ readings. Patients having ‘satisfactory clinical condition’ in Day-1 and 5 are more likely to have a stable or satisfactory outcome. However, this feature is less prominent in critical/serious group’s decision rules. (Figure 2)

Based on the testing dataset, the current XGboost classifier (“the full model”) achieved a very high predictive accuracy rate of 92% and 99% to classify study subjects into three outcome groups on day 1 and day 5 of admission, which is on average equivalent to day 6 and day 10 of symptom onset given the median/mean onset-to-admission duration being 4/5.8 days. In this big data era, it’s technically feasible to automate predicted probabilities of three outcome classes as predicted from this methodology which simultaneously considers a multitude of factors readily available in HA’s CMS, and is compatible with missing data values in study subjects, commonly encountered when using observational data at service settings.

In order to provide quick and accurate prediction at the point of care, simplication of the predictive modelling is suggested. Through a robust variable selection process of the two full models on day 1 and day 5 of admission, a simpler model with seven common specific predictors were identified. They included the worst clinical condition status (4-level), age group, and five biomarkers, namely, CRP, LDH, platelet, N/L ratio and A/G ratio. By using these modified models at the point of care, physicians will have additional and convenient information on the prognostic analysis of the COVID-19 patients at early presentation to hospitals. This will increase the turnaround time of our precious inpatient facilities and thus the efficiency of infection control measures.

Experts worldwide warn that COVID-19 may persist into this winter. For Hong Kong to continue with the current “early identification, early isolation and early treatment” strategy, this prognostic modelling is part of the corporate strategy in the preparedness for the potential third wave of outbreak in Hong Kong. Learning from the examples in nearby countries, like Singapore, Hong Kong is preparing the emergency response in case over hundreds or thousands of new cases presented per day, which will overwhelm our existing isolation bed capacity in the public hospital system. Planning for community isolation and treatment facilities is underway. In order to ensure patient safety and quality care, a tool to identify patients with lesser severity and better outcome is necessary. This group of patients probably will be safely managed at the community settings. This study provides a robust analysis on prognostication for COVID-19 cases. The transformation into a simple calculator tool to predict clinical outcome on day 1 and day 5 of admission makes it more convenient for clinicians to apply in their daily settings. It allows early identification of newly confirmed cases upon presentation and triage to appropriate places for isolation and treatment at appropriate timing.

Limitation of the study includes the lack of inclusion of data from radiological imaging. Patients with COVID-19 are found to have lung infection with ground glass and consolidative opacities with peripheral and lower lung distribution and bilateral involvement [29]. We are going to include all chest X-ray images up to day 5 in the next study through AI approach of image analytics.

## Conclusion

This study provides comprehensive analysis on epidemiological, clinical and laboratory data within the first five days of admission among the entire Hong Kong cohort of 1,037 laboratory-confirmed COVID-19 patients. Having considered a multitude of prognostic factors, the model could accurately predict the clinical outcome of a COVID-19 case on day 1 and day 5 of admission, namely, critical/serious, stable and satisfactory. It aims to serve as a management tool as well as a clinical reference tool in order to plan ahead for response measures on triage and step-down care to cope with the next unprecedented wave of COVID-19 epidemic. With a trade-off between practical application and predictive accuracy, the full model consisting of 30 features were reduced into a simpler model with seven specific features in tandem with a simple, easy-to-understand and transparent calculator tool for applications locally and open access globally.

## Data Availability

The de-identified datasets generated and analysed during the current study are not publicly available for patient privacy protection as their disclosure at granular level may entail the risk of subject re-identification. Aggregate data are available from the corresponding author on reasonable request.
The calculator tool which is developed based on the prognostic models' results of this study will be available for online public access after the study being published.

## List of Abbreviations

A: albumin
A/G: ratio albumin-globulin ratio
AI: Artificial Intelligence
AII: airborne infection isolation
ALP: alkaline phosphatase
ALT: alanine aminotransferase
AST: aspartate aminotransferase
CMS: Clinical Management System
COVID-19: Coronavirus Disease 2019
CRP: C-reactive protein
ECMO: extracorporeal membrane oxygenation
eNID: Electronic Notification of Infectious Disease
G: globulin
HA: Hospital Authority
HK: Hong Kong
ICU: intensive care unit
ILI: influenza-like illness
LDH: lactate dehydrogenase
MPV: mean platelet volume
N/L: ratio neutrophil-lymphocyte ratio
NDORS: Notifiable Diseases and Outbreak Reporting System
PCT: procalicitonin
RT-PCR: reverse-transcription polymerase chain reaction
SARS: Severe Acute Respiratory Syndrome
SD: standard deviation
WBC: white blood cell count
WHO: World Health Organization
XGBoost: Extreme Gradient Boosting

## Appendix.

**Appendix Figure 1.**
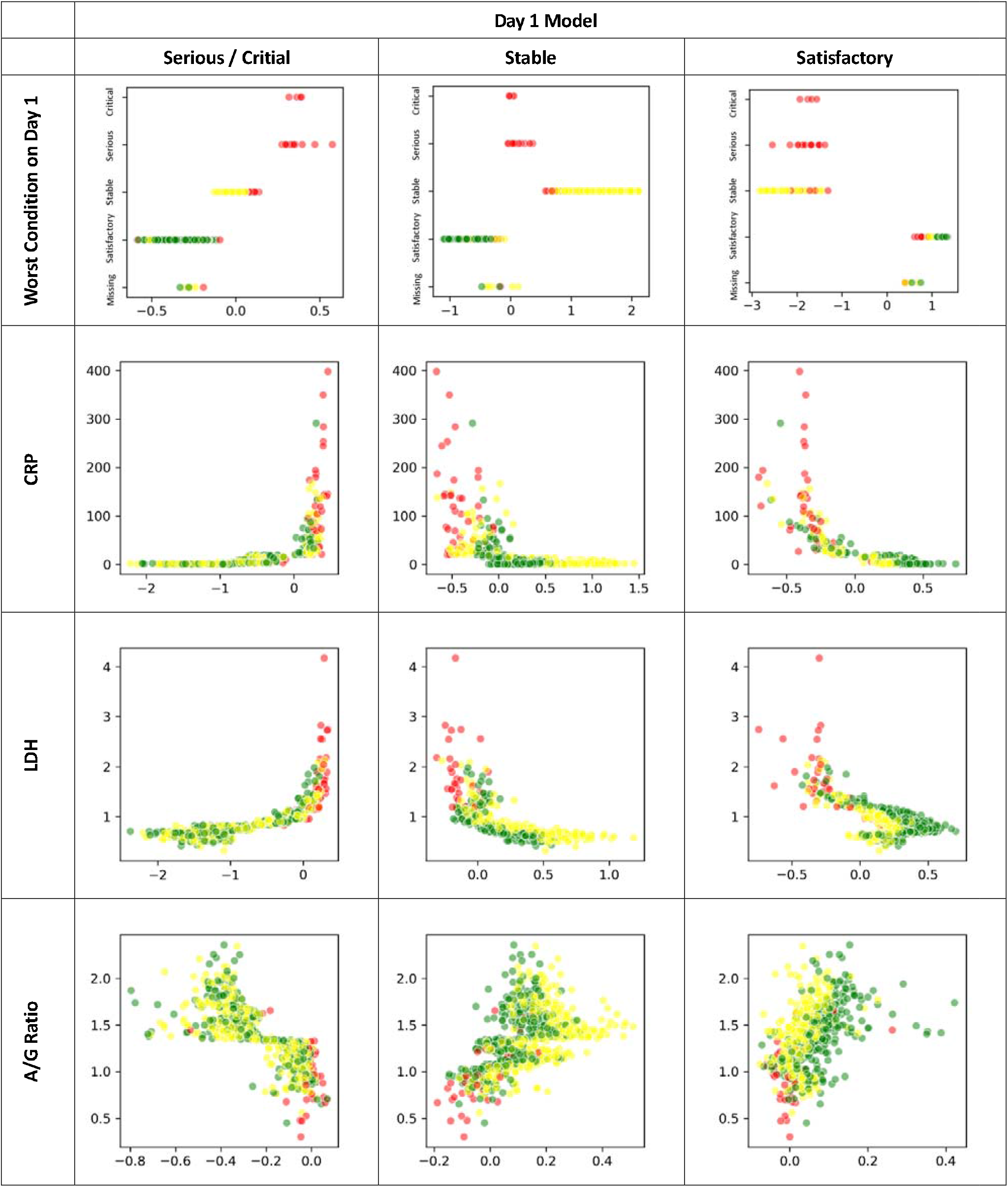
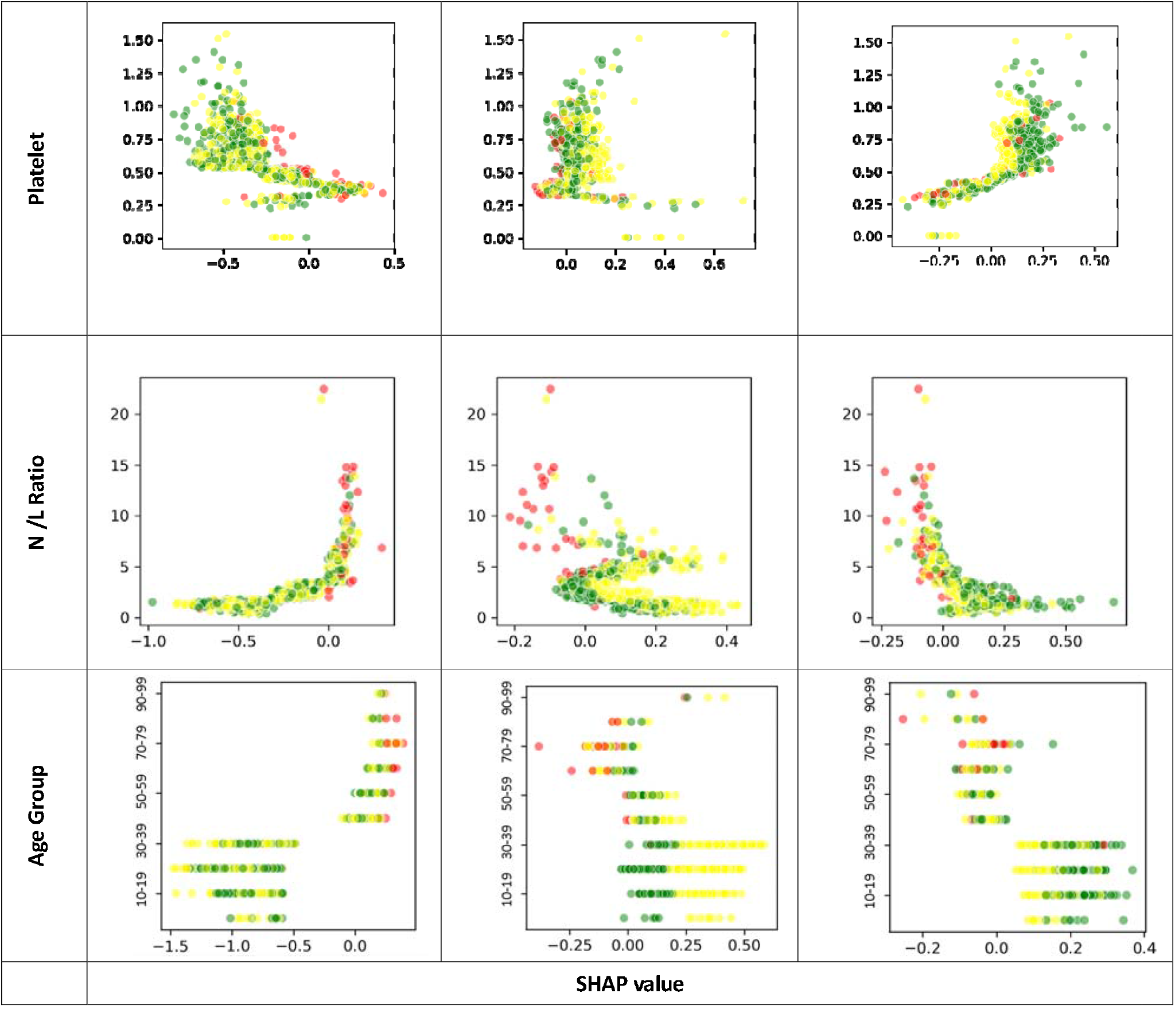
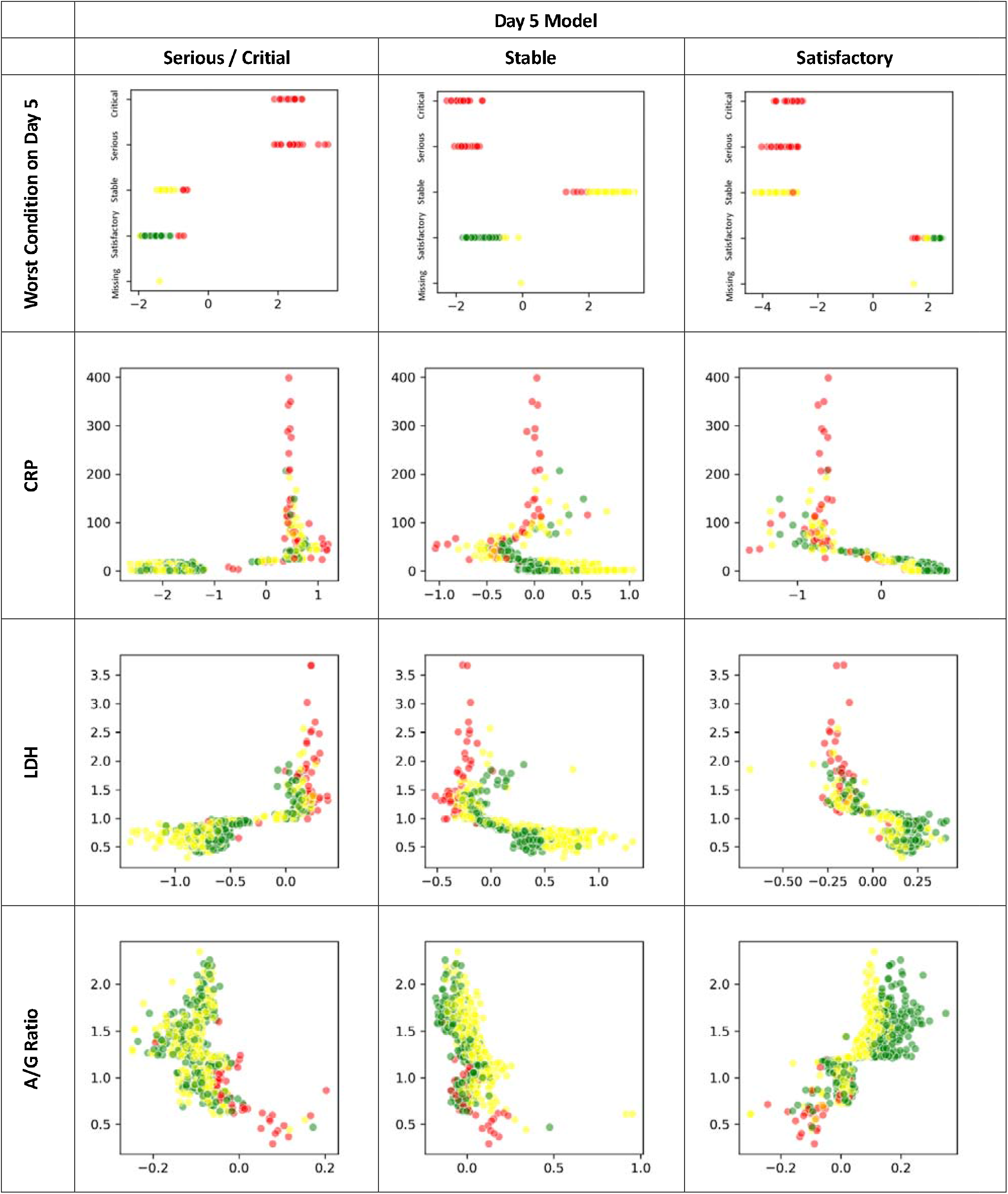
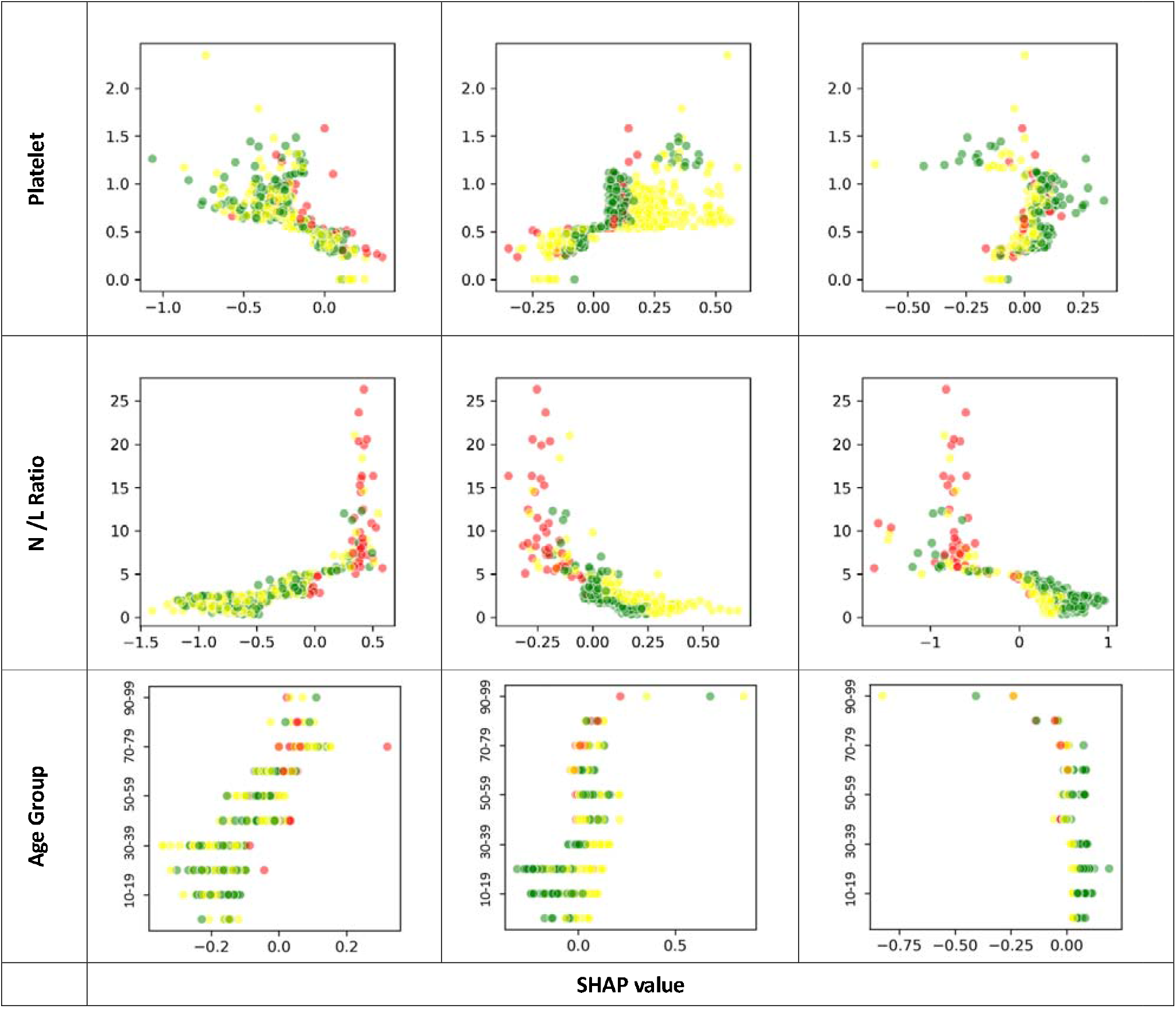
Relationship between each key feature under Day 1 and Day 5 of admission and the SHAP value for each outcome group (Red: Critical / Serious, Yellow: Stable, Green: Satisfactory)

## References

1. WHO Coronavirus Disease (COVID-19) Dashboard. World Health Organization. 2020. https://covid19.who.int/. Accessed 20 May 2020.

2. Latest situation of cases of COVID-19 (as of 11 May 2020). Centre for Health Protection, HKSAR. 2020. https://www.coronavirus.gov.hk/eng/index.html. Accessed 20 May 2020.

3. Gibney E. Whose coronavirus strategy worked best? Scientists hunt most effective policies. Nature. 2020;581:15–16.

4. Countries by density 2020. World population review. 2020. https://worldpopulationreview.com/countries/countries-by-density/. Accessed 20 May 2020.

5. Introduction. Hospital Authority, HKSAR. 2020. https://www.ha.org.hk/visitor/ha_visitor_index.asp?Content_ID=10008&Lang=ENG&Dimension=100&Parent_ID=10004&Ver=HTML. Accessed 20 May 2020.

6. Cheung N-T, Fung V, Kong JHB. The Hong Kong Hospital Authority’s information architecture. Stud Health Technol Inform. 2004;107(2):1183–6.

7. SCMP Editorial. We can all help ease burden on hospitals. South China Morning Post. 2020. https://www.scmp.com/comment/opinion/article/3077795/we-can-all-help-ease-burden-hospitals. Accessed 20 May 2020.

8. HA Task Force on Clinical Management on Infection. Interim Recommendation on Clinical Management of Adult Cases with Coronavirus Disease 2019 (COVID-19). Hospital Authority, HKSAR; 2020. https://ha.home/ho/cico/Interim_Recommendation_on_Clinical_Management_of_Adult_Cases_with_COVID-19.pdf. Accessed 20 May 2020.

9. HA Task Force on Clinical Management on Infection. Interim Recommendation on Clinical Management of Paediatric Patients of Coronavirus Disease 2019 (COVID 19) Infection. Hospital Authority, HKSAR; 2020. https://ha.home/ho/cico/Interim_Recommendation_on_Clinical_Management_of_Paediatric_Patients_of_COVID-19infection.pdf. Accessed 20 May 2020.

10. World Health Organization. Global surveillance for COVID-19 caused by human infection with COVID-19 virus: interim guidance. 2020. https://www.who.int/publications-detail/global-surveillance-for-covid-19-caused-by-human-infection-with-covid-19-virus-interim-guidance. Accessed 20 May 2020.

11. Hong Kong Cancer Registry. Hospital Authority, HKSAR. https://www3.ha.org.hk/cancereg/. Accessed 20 May 2020.

12. Chen R, Liang W, Jiang M, Guan W, Zhan C, Wang T, et al. Risk Factors of Fatal Outcome in Hospitalized Subjects With Coronavirus Disease 2019 From a Nationwide Analysis in China. CHEST. 2020; doi:10.1016/j.chest.2020.04.010.

13. Zhang J, Wang X, Jia X, Li J, Hu K, Chen G, et al. Risk factors for disease severity, unimprovement, and mortality in COVID-19 patients in Wuhan, China. Clinical Microbiology and Infection. 2020;26(6):767–772.

14. Zhang L, Yan X, Fan Q, Liu H, Liu X, Liu Z, et al. D-dimer levels on admission to predict in-hospital mortality in patients with Covid-19. J Thromb Haemost. 2020;18(6):1324–1329.

15. McIntosh K. Coronavirus disease 2019 (COVID-19): Epidemiology, virology, clinical features, diagnosis, and prevention. UpToDate. https://www.uptodate.com/contents/coronavirus-disease-2019-covid-19-epidemiology-virology-clinical-features-diagnosis-and-prevention?sectionName=Strategies%20for%20PPE%20shortages&topicRef=127556&anchor=H775838145&source=see_link#H3392906512. Accessed 19 May 2020.

16. Zhou F, Yu T, Du R, Fan G, Liu Y, Liu Z, et al. Clinical course and risk factors for mortality of adult inpatients with COVID-19 in Wuhan, China: a retrospective cohort study. The Lancet. 2020;395:1054–62.

17. Zheng Z, Peng F, Xu B, Zhao J, Liu H, Peng J, et al. Risk factors of critical & mortal COVID-19 cases: A systematic literature review and meta-analysis. Journal of Infection. 2020; doi:10.1016/j.jinf.2020.04.021.

18. Zhu Z, Cai T, Fan L, Lou K, Hua X, Huang Z, et al. Clinical value of immune-inflammatory parameters to assess the severity of coronavirus disease 2019. International Journal of Infectious Diseases. 2020;95:332–9.

19. Yao Q, Wang P, Wang X, Qie G, Meng M, Tong X, et al. Retrospective study of risk factors for severe SARS-Cov-2 infections in hospitalized adult patients. Pol Arch Intern Med. 2020;130(5):390–399.

20. Liang W, Liang H, Ou L, Chen B, Chen A, Li C, et al. Development and Validation of a Clinical Risk Score to Predict the Occurrence of Critical Illness in Hospitalized Patients With COVID-19. JAMA Intern Med. 2020; doi: 10.1001/jamainternmed.2020.2033.

21. Yan L, Zhang H-T, Goncalves J, Xiao Y, Wang M, Guo Y, et al. An interpretable mortality prediction model for COVID-19 patients. Nature Machine Intelligence. 2020;2:283–8.

22. Cummings MJ, Baldwin MR, Abrams D, Jacobson SD, Meyer BJ, Balough EM, et al. Epidemiology, clinical course, and outcomes of critically ill adults with COVID-19 in New York City: a prospective cohort study. The Lancet. 2020;395:1763–70.

23. Qu R, Ling Y, Zhang Y, Wei L, Chen X, Li X, et al. Platelet-to-lymphocyte ratio is associated with prognosis in patients with coronavirus disease-19. J Med Virol. 2020; doi: 10.1002/jmv.25767.

24. Chan JCK, Tsui ELH, Wong VCW, Hospital Authority SARS Collaborative Group. Prognostication in severe acute respiratory syndrome: a retrospective time-course analysis of 1312 laboratory-confirmed patients in Hong Kong. Respirology. 2007;12:531–42.

25. Tsui PT, Kwok ML, Yuen H, Lai ST. Severe Acute Respiratory Syndrome: Clinical Outcome and Prognostic Correlates1. Emerg Infect Dis. 2003;9:1064–9.

26. Choi KW, Chau TN, Tsang O, Tso E, Chiu MC, Tong WL, et al. Outcomes and prognostic factors in 267 patients with severe acute respiratory syndrome in Hong Kong. Ann Intern Med. 2003;139:715–23.

27. Fushiki T. Estimation of prediction error by using K-fold cross-validation. Stat Comput. 2011;21:137–46.

28. National Health Commission of the People’s Republic of China. Diagnosis and Treatment Protocol for COVID-19 (Trial Version 7). 2020. http://en.nhc.gov.cn/2020-03/29/c_78469.htm. Accessed 20 May 2020.

29. Wong HYF, Lam HYS, Fong AH-T, Leung ST, Chin TW-Y, Lo CSY, et al. Frequency and Distribution of Chest Radiographic Findings in COVID-19 Positive Patients. Radiology. 2019;201160.

